# Co-development of a digital animated video on ADHD with children and families/carers

**DOI:** 10.1101/2025.08.18.25333941

**Authors:** Sharifah Shameem Agha, Rhys Bevan-Jones, Lowri O’Donovan, Sarah-Jane Bailey, Catrin Hopkins, Anita Thapar, Kate Langley

## Abstract

**Background:** When receiving an attention deficit hyperactivity disorder (ADHD) diagnosis, children and families/carers need clear and accurate information about the condition in a suitable format. However, many of the available resources are complex or provide content that is not evidence-based. Furthermore, there are limited studies that co-develop digital resources with and for children, particularly those with ADHD. This study describes a co-production approach with an under-represented group, children with ADHD (aged 7-11), and their families/carers, to develop an animated video to provide support and improve understanding of ADHD.

**Methods:** The iterative co-design process involved a series of focus groups with children and families/carers at different stages of video animation development. The views of healthcare professionals were also obtained via online questionnaires and interviews. Key themes were identified through thematic analysis before more detailed discussions of content and a review of the animation storyboard.

**Results:** Eleven families (12 children, 11 parents/carers) were involved in focus groups and 23 health professionals responded to online questionnaires. The study resulted in the creation of a widely accessible evidence-based bilingual (English, Welsh) animated video for children aged 7-11, newly diagnosed with ADHD, and their families/carers. Preliminary feedback and evaluation from participants at the animation launch event showed that the animation has been well received.

**Conclusions:** This study provides background around the development of a well-received digital resource for children with ADHD. It also outlines a framework for the co-production of further resources, especially as those involved agreed that a wider range of resources is needed for children and young people with ADHD.

## Introduction

Attention Deficit Hyperactivity Disorder (ADHD) is a neurodevelopmental condition characterized by impairing levels of concentration, overactivity and impulsivity that impacts individuals and their families (Thapar & Cooper, 2016). Despite affecting 3-5% of children (Thapar & Cooper, 2016), ADHD is poorly understood and associated with much stigma (Bisset et al., 2021). Children and parents/carers who are undergoing a neurodevelopmental assessment may not have a full understanding of ADHD (DosReis et al., 2010; McKenna et al., 2024). Also, growing demands on clinicians’ time mean there is little time for psychoeducation or post diagnostic support as recommended by NICE (NICE, 2019). Thus, initial diagnosis can be a difficult time when children with ADHD and their families/carers are faced with challenges, especially when seeking out information or resources, including those explaining the condition to children (Hamed et al., 2015; McKenna et al., 2024; Wright et al., 2015).

Although there are effective interventions (pharmacological and nonpharmacological) for ADHD, it is reported that the use and adherence to treatment are low, resulting in suboptimal outcomes for some individuals (Baweja et al., 2021; Corkum et al., 2015). Research has identified several barriers to engagement with interventions, including lack of knowledge, stigma and negative beliefs about ADHD (Baweja et al., 2021). One method for overcoming these barriers is through psychoeducational tools (Baweja et al., 2021; Dahl et al., 2020).

Psychoeducation can be defined as a systematic and didactic method of informing individuals and their families/carers about their condition and support to help with understanding and management (Ferrin et al., 2014). The National Institute for Health and Care Excellence (NICE, 2019) recommend psychoeducation as the first line of management for ADHD, emphasising the importance of providing information and support to individuals and parents/carers following a diagnosis. Studies have shown that psychoeducation in ADHD can lead to improvement in engagement in services and improvement of ADHD symptoms (Bai et al., 2015; Corkum et al., 1999; Dahl et al., 2020; Ferrin et al., 2014; Morris et al., 2025).

Despite emphasis on the value of psychoeducation, families/carers, support groups and researchers have reported that there are insufficient resources available. Further investigation has suggested that many available resources are not fit for purpose, are rigid, not tailored to individual’s needs, are too complex for children, or provide content that is not evidence-based and is of poor quality (King et al., 2021; Montoya et al., 2013; Ward et al., 2020; Yeung et al., 2022; Young et al., 2021). Much of the available information is aimed at parents and carers rather than children (Lantz et al., 2021; MacKay & Corkum, 2006; Morris et al., 2025; Young et al., 2021) or are family-based interventions (Schoenfelder et al., 2020). A review exploring the unmet needs of those with ADHD has highlighted that there is a lack of in depth, supportive psychoeducational resources free from negative narratives (Bisset et al., 2023). Experts also emphasised the need for high-quality, age and presentation-appropriate psychoeducation for those with a new diagnosis (Young et al., 2021).

Digital tools and platforms can be effective in improving mental health literacy and improving stigmatising attitudes (Ito-Jaeger et al., 2022; Yeo et al., 2024). The internet and social media have been reported to be the preferred source of information for ADHD for both adolescents and parents (Bussing et al., 2012a; Sciutto, 2015). However, information and advice found on digital media can be misleading and inaccurate. Studies investigating the quality of online information and videos (e.g. YouTube and TikTok) on ADHD have found overall that the quality of information was poor, misleading and were from unqualified creators (Montoya et al., 2013; Ward et al., 2020; Yeung et al., 2022). Therefore, having clear and accurate information in a digital format might help children to understand and manage their condition.

To ensure resources are relevant, acceptable and meet user needs and preferences, it is important to engage potential users and stakeholders in their development and production (Bevan Jones et al., 2020; Bisset et al., 2023). There are some published examples of the co-development of resources for young children with ADHD (Powell et al., 2021) (Armitt et al., 2022) but this is limited. Individuals with ADHD are largely excluded from the research and design of products and resources (Spiel et al., 2022).

Following initial discussions around the need to counter the lack of suitable co-produced resources we aimed to co-produce an animation video with children aged 7-11 years with ADHD and parents/carers, which provides information and support to those with a diagnosis.

## Methods

Researchers and clinicians at Cardiff University and Cwm Taf Morgannwg University Health Board (UHB) worked in partnership with a local ADHD parent support group charity (ADHD Cardiff). Ideas for this project started out as general discussions with SJ (charity trustee ADHD Cardiff). Input from stakeholders, including children with ADHD, families/carers and practitioners, helped steer the project and they were engaged in every stage of the project from conceptualisation to the launch of the final product. The project had four stages: 1) Development of ideas, 2) Content development and review 3) Development of script and storyboard, 4) Animation launch and initial evaluation (see Figure 1).

**Figure 1:**
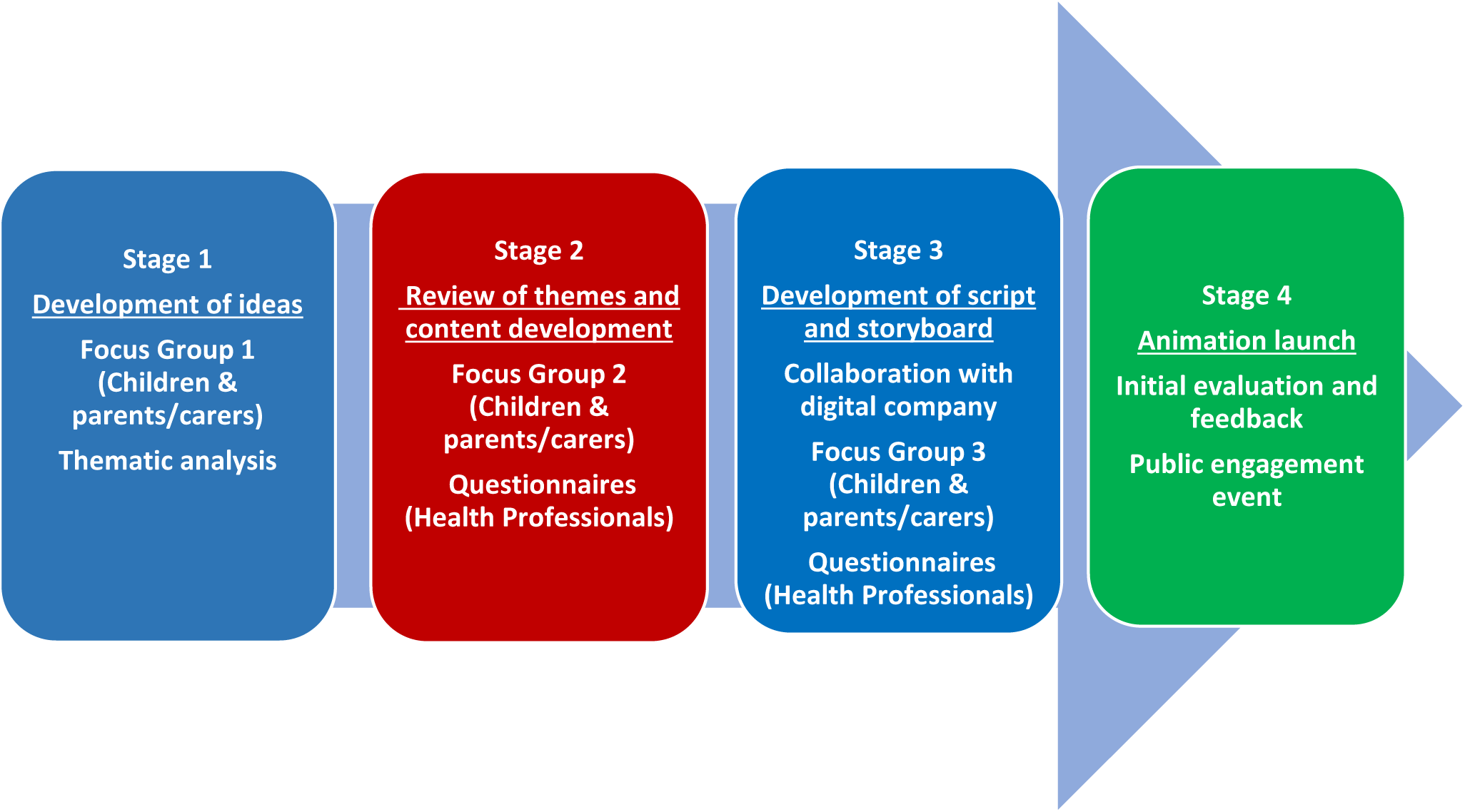
Stages of the video development process.

### Participants

#### Children with ADHD and parents/carers

Children with a diagnosis of ADHD (aged 7-11 years) and their parents/carers were invited through advertisement via closed social media network linked with ADHD Cardiff. There were no exclusion criteria based on child or parent/carer characteristics or ADHD profile. Informed consent was obtained from parents/carers and informed assent was obtained from the child to take part in the project.

#### Health professionals

Health professionals from secondary care who work with children or young people with ADHD were invited to take part in the study. They were identified through existing researcher networks, and informed consent was obtained.

Ethical approval for the study was obtained from the School of Medicine, Cardiff University (SMREC 18/42).

### Focus groups and questionnaires

The animation video was developed and co-produced in a staged iterative process through a series of focus groups (FGs) with children and with parents/carers. Six FGs in total were undertaken, three each for children and parents/carers at stages 1, 2 and 3 of the development process (see Figure 1).

The FGs drew upon principles of the Diversity for Design Framework (D4D) for ADHD (Fekete & Lucero, 2019) whereby sessions were tailored to suit participants and the environment structured to create a safe and comfortable space for everyone to engage. The D4D framework emphasises maximising children’s strengths and supporting potential difficulties (Fekete & Lucero, 2019). To ensure meaningful participation, the FGs were carefully planned after consultation and discussion with potential participants and a charity trustee (SJ). All the FGs were organised after school hours and were facilitated by KL (parent/carer FG) and by SSA (children FG), and supported by other researchers (including RBJ & LOD). Upon arrival, children and families/carers were provided with refreshments.

During the FG sessions, multiple modes of expression, appropriate content, and visual and task-based activities were prepared to ensure that everyone had a voice and opportunity to contribute (see details below). Each parent focus group was 1.5-2 hours in duration. The children had shorter, interactive sessions (30 minutes) followed by engaging workshops (circus skills, learning magic tricks and beatboxing) whilst their parents/carers finished their session. FGs and workshops were held at Cardiff University seminar rooms and exhibition space. A quiet room was provided for participants if they needed a break or a safe space to relax. There was also an activity area for siblings who accompanied parents/carers but did not participate in the co-production activities. FGs were audio recorded and professionally transcribed. Participants could also write or draw their ideas and thoughts.

Throughout the process, views of health professionals were obtained and integrated via meetings with the first author (SSA) and online questionnaires. They were consulted at two separate stages of the development process: Stage 2 - review of preliminary themes generated from the first focus groups and Stage 3 - review of the storyboard draft for the video, concurrent with the final focus groups.

### Development process

#### Stage 1: Development of ideas

The first FG aimed to generate ideas from participants about the potential content and areas that should be addressed in the prospective video. Discussions were held in the FGs using the topic guide which was informed by the project aims. Questions included participants’ experiences of ADHD before and after diagnosis, their experience of looking for information about ADHD and their views on what should be included in the video. Researchers presented “myths and facts” about ADHD, as a starting point for gathering ideas for the video content. These “myths and facts” have been used by the researchers in previous public engagement work and feedback demonstrated that this provides an engaging and useful basis for communicating an understanding of ADHD with a scientific evidence-based foundation. Examples discussed were ‘ADHD is not a new condition, ADHD is not caused by parenting or junk food, ADHD is not just a condition for children’ (Topic guides - Appendices 1a & 2a). All the information presented at the FGs were conducted via presentation slides and posters.

#### Analysis

The data from the FG was analysed using an inductive thematic analysis approach. The analysis followed Braun & Clarke’s six-phase framework for thematic analysis; familiarity with data, generating initial codes, searching for themes, reviewing themes, defining and naming themes and writing up) (Braun & Clarke, 2006). Analysis was also conducted as an iterative process where themes generated were fed into the stage 2 co-production activities for review and refinement (FG 2 and health professional questionnaire). The transcripts from the first stage of FGs were coded independently by SSA and LOD. Initial agreement on coding and themes was discussed to obtain consensus and subsequently with other authors before presenting the suggested preliminary themes to stakeholders. Codes were categorised into subcodes and collated into preliminary key themes which helped to inform the content for stage 2. Thematic analysis was conducted using qualitative analysis software NVivo (QSR International) (version 12).

#### Stage 2: content development and review

The aim of the second FG was to review themes from stage 1 and further develop visual ideas. Before the second FG session, parent/carers were sent a summary of the preliminary themes so they could reflect on and discuss them with their children. The themes were discussed in the child FG through simple presentation slides. RBJ developed illustrations based on children’s ideas to stimulate further discussions. At the FG, parents/carers completed an individual questionnaire to rate the importance of each theme and which issues to avoid. Group discussions about each preliminary theme then used the questionnaire responses as a guide.

Around the same time as FG2, health professionals were sent an online questionnaire to review the preliminary themes and were asked to rate each theme using similar questions as were asked in the parent/carer groups (Appendix 3a).

#### Analysis

The preliminary themes were reviewed and refined further to ensure it reflected findings obtained from stakeholder discussion. Each was verified with references from credible peer-reviewed sources and knowledge and expertise of the research team to ensure accuracy of content.

#### Stage 3: Development of script and storyboard

Once the key themes were refined, SSA wrote the draft animation script based on the themes and in line with the research evidence on ADHD. RBJ developed initial visual designs and ideas based on suggestions from the groups. These were reviewed by other co-authors (KL, AT). The script was also translated into Welsh to ensure inclusivity for those in Wales and in accordance with NICE guidelines that children and families/carers should be communicated with in their first language where possible (NICE 2021). The research team then worked closely with a digital media company to create an animation storyboard.

In the third round of FGs, participants were asked to provide feedback on the storyboard, script, designs of the visual ideas and characters, and identified whether the content themes had been adequately addressed (Appendix 1c & 2c). The storyboard was also printed out as posters (Figure 2), and parents/carers were asked to stick post-it notes on these with their thoughts and comments. For the child FG, participants were split into groups of 2 or 3 children accompanied by facilitators and were asked to put different stickers on parts of the storyboard that they liked or did not like.

**Figure 2:**
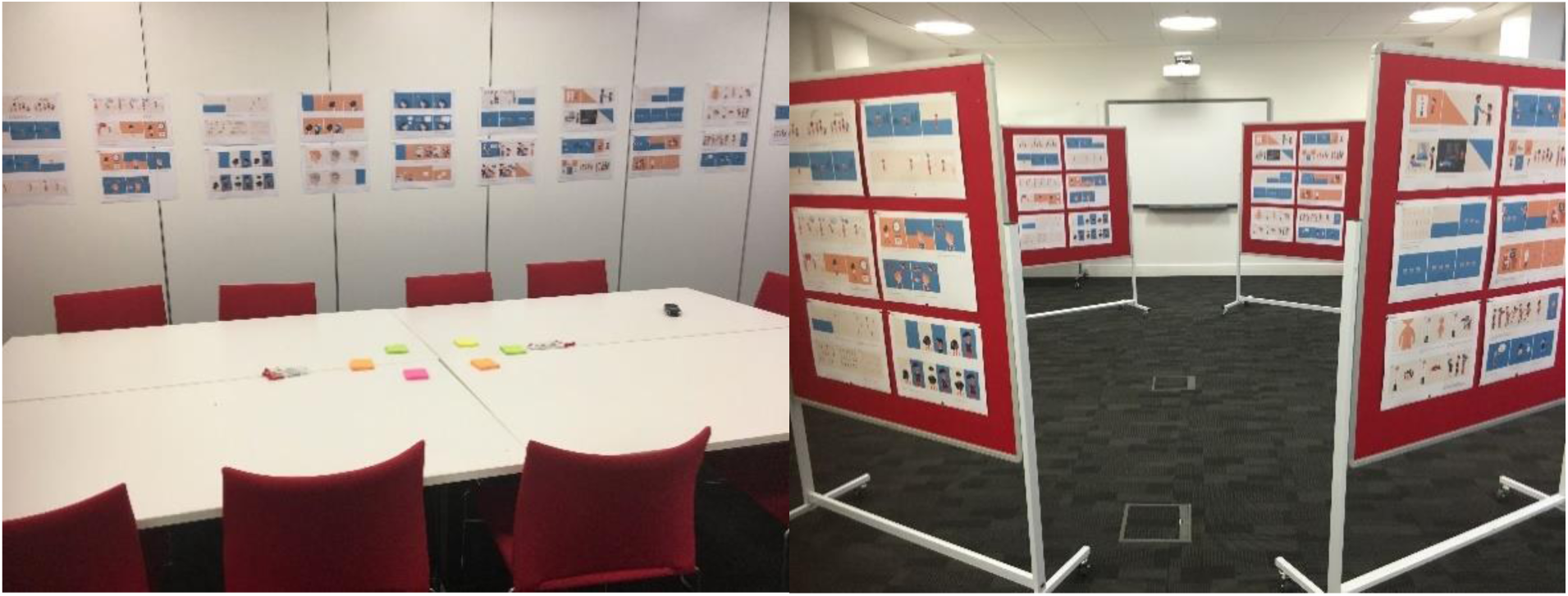
Parent/carer (left) & Child (right) focus group set up.

Health professionals were sent a copy of the storyboard and an online questionnaire on their thoughts on the storyboard, for examples whether key themes were adequately addressed, and the messages portrayed were appropriate or well-described (see Appendix 3b).

#### Stage 4: Animation launch and evaluation

Changes were made based on feedback received in FG 3 and the final ADHD video animation was produced. The voiceovers for the animation were bilingual (English and Welsh) and chosen from a selection of child voiceover actors. Children from the FG were also given the opportunity to record sections of the voiceovers. Separate written permission and consent were obtained for this.

To celebrate and show appreciation for the work of all those involved, a launch and public engagement event was held for key stakeholders. Focus group participants, members of the ADHD parent support groups, and healthcare/educational professionals from across South Wales were invited to attend. Activities included workshops, brain games activities (which included conducting science experiments), talks on our latest ADHD research and premiere of the animation video. All participating children were awarded a certificate for their contribution. At the event, parents/carers and professionals were given a questionnaire about their opinions of the animation, whether the animation would be useful for children who have been diagnosed and suggestions for the animation title. Children were asked to rate the animation video on a poster using stickers and post-it notes.

## Results

### Focus group participants

Eleven families (12 children, 11 parents/carers) took part in the first FG. All were invited back for the subsequent sessions, and seven families attended the second and third groups (retention rate of 64%). Mean child age at stage 1 was 9.2 years (SD 1.66). Nine children (75%) were male (parent-reported). All parents/carers who attended the focus groups were female.

### Summary of themes from child and parent/carer FGs (stage 1)

Six key themes were identified from FG 1 (Table 1). The first two reflected practical considerations: 1) Importance and need for resources and 2) video design features. Four additional themes were used to form the main content of the animation storyboard: 3) Information about ADHD, 4) Self-management tips, 5) Positive aspects of ADHD, and 6) Challenges of ADHD. The key themes and subthemes on animation content derived from the FG are presented in Table 1. The themes and subthemes were endorsed by all participants, through quotes which are presented in table 2.

**Table 1:** Structure of themes and subthemes.

| Theme category | Themes | Subthemes from focus groups | Subthemes from questionnaires |
| --- | --- | --- | --- |
| Themes on practical considerations | 1.Importance and need for resource | Lack of resources<br>Current material unsuitable<br>Important to educate | Lack of accessible resources readily available<br>Useful to raise awareness |
|  | 2. Video design features | Subtitles<br>Use of children's voices<br>Visual ideas to describe science<br>Animated | Visual support aids or examples |
| Content themes | 3. Information about ADHD | Understanding ADHD<br>Information about diagnosis<br>Facts and figures<br>Diversity | Understanding ADHD<br>Educating families<br>Increasing awareness and reducing stigma<br>Diversity |
|  | 4. Self-management tips | Helpful tips<br>Managing symptoms<br>Medication | Self-management is important<br>Empowering families<br>Supportive and practical advice<br>Medication not only solution<br>Individual differences |
|  | 5. Positive aspects of ADHD | Positive messages<br>Strengths / positive ADHD traits<br>Future<br>Celebrities<br>Negative associations with ADHD<br>Helping with self esteem | Negative stereotype<br>Self-esteem<br>Positive of ADHD<br>Advantages/strengths |
|  | 6. Challenges of having ADHD | Having ADHD is hard work<br>Co-occurring difficulties<br>Friendships and social communication | Understanding difficulties<br>Recognising struggles<br>Understanding limitations |

**Table 2:**
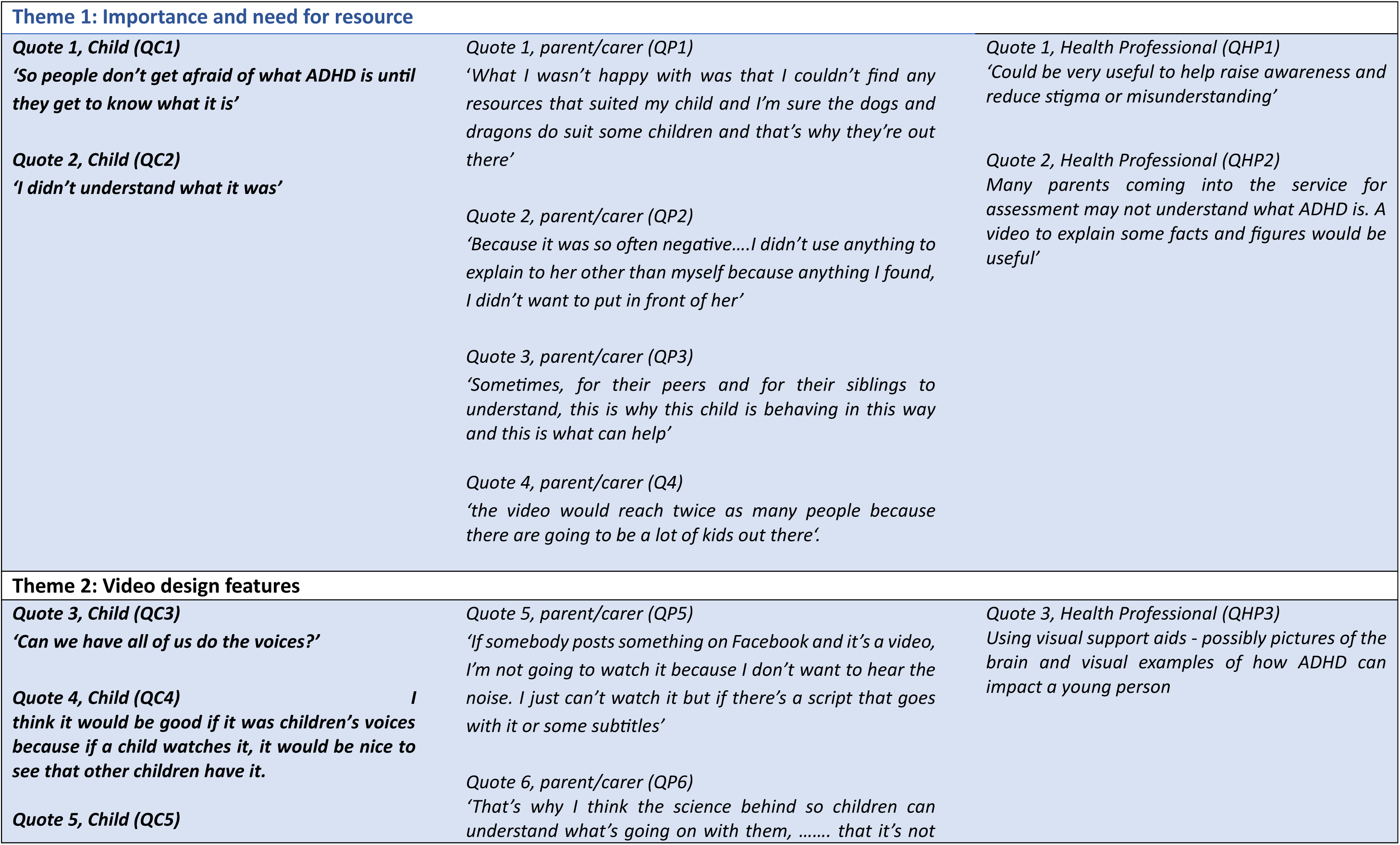
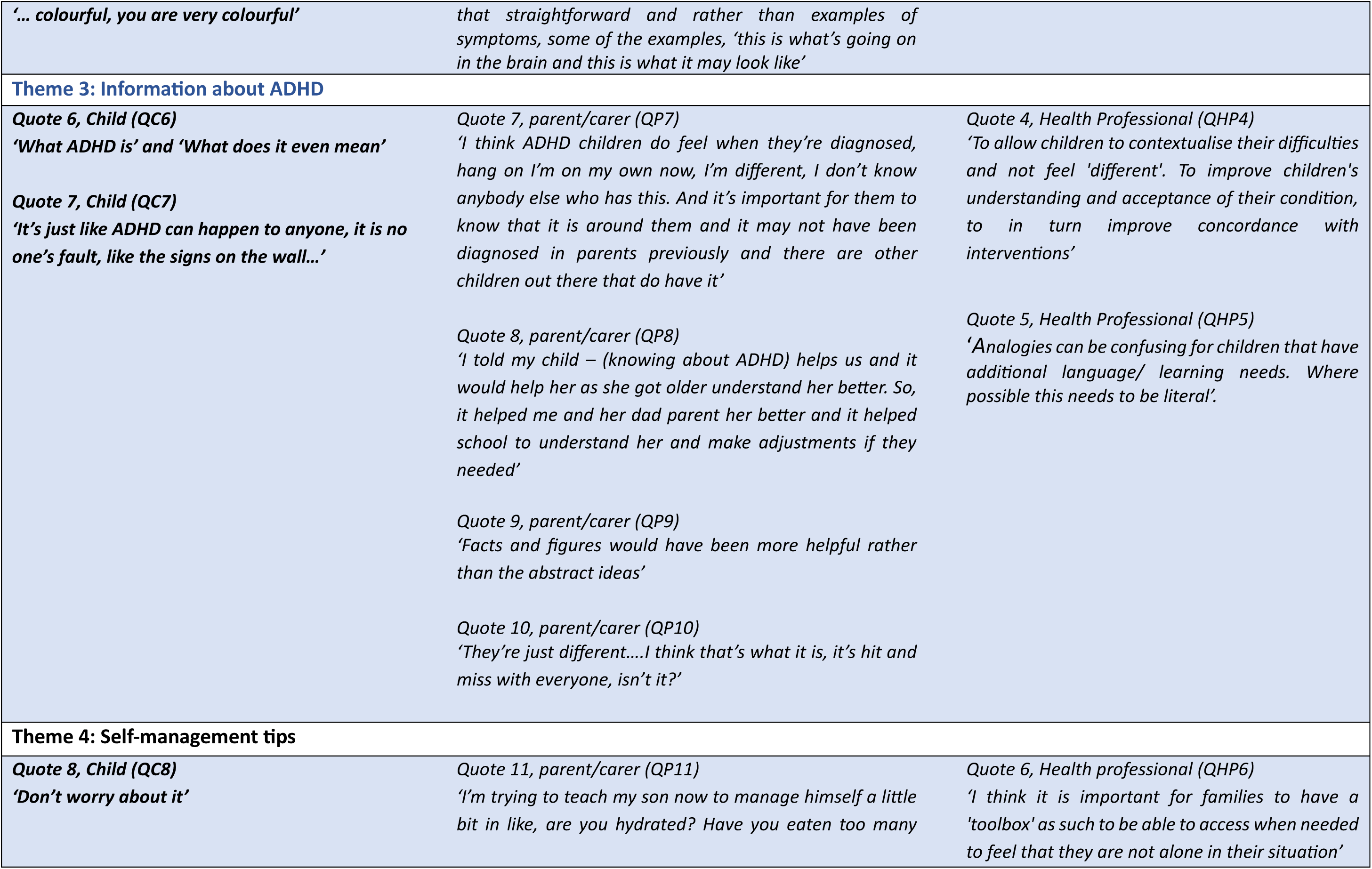
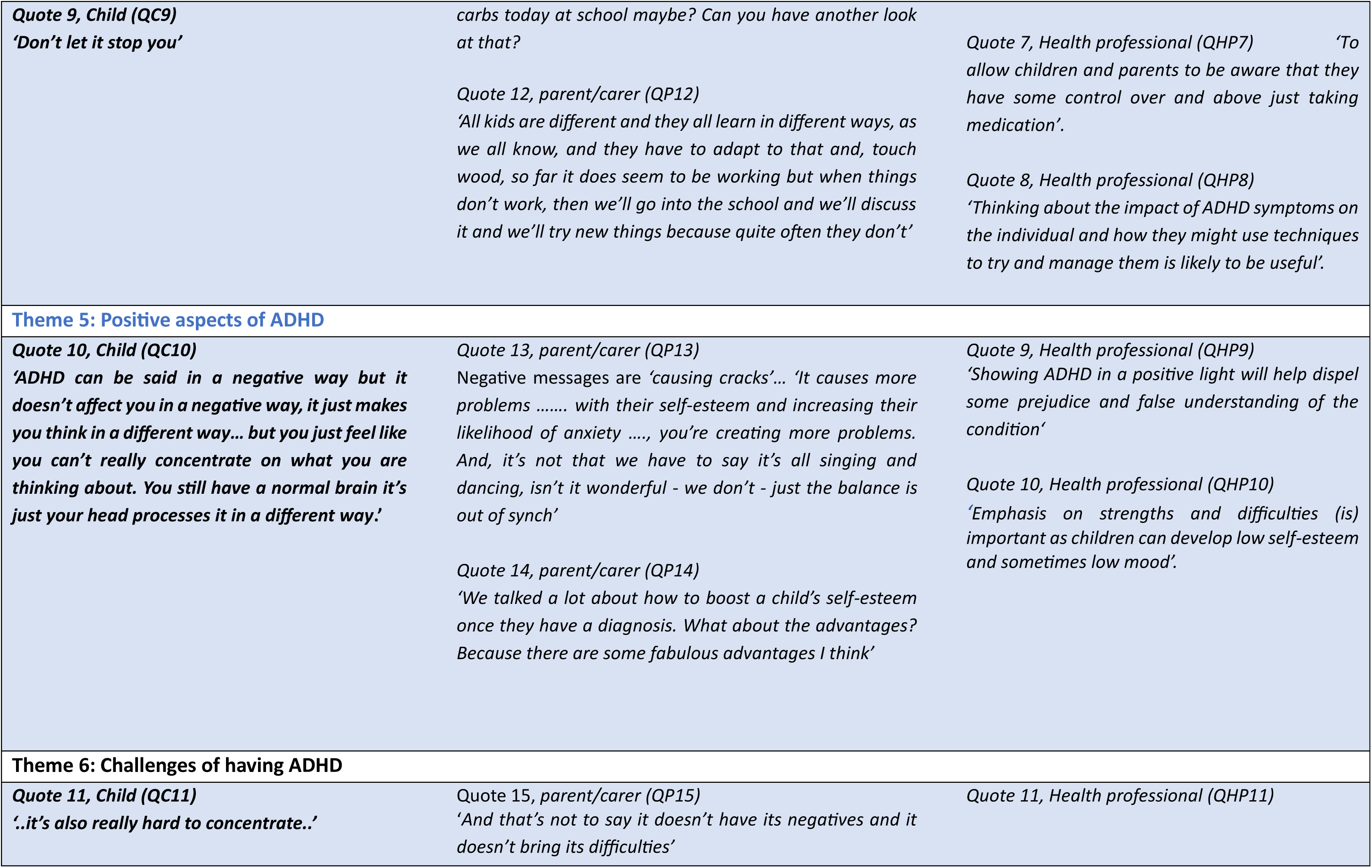
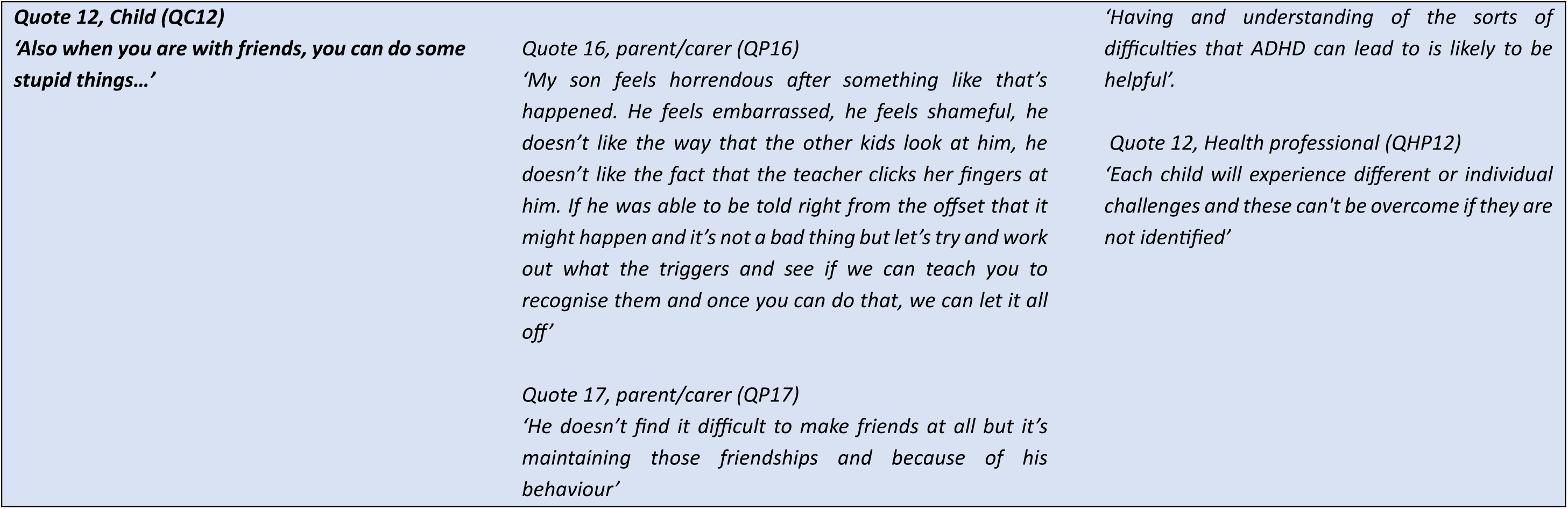
Quotes supporting themes and subthemes.

#### · Theme 1: Importance and need for resources

All FG participants agreed that there is a lack of resources or materials to explain what ADHD is to children (quote 1, parent/carer (QP1), table 2). Children felt that a video resource could reduce initial fear of the condition (quote 1, Child (QC1) in table 2). This was also reflected in their own lack of understanding when first diagnosed with ADHD (QC2). Parents/carers found that what was already available was not suitable or appropriate (QP1; QP2). Participants also highlighted the need to educate others like peers, siblings and family members, so they have a better understanding of ADHD (QP3). Furthermore, presenting information in a video format would be beneficial as it would improve reach (QP4).

#### · Theme 2: Video design features

Participants had discussed what features the video should include. Parent/carer suggestions included the use of subtitles and children’s voices (QP5). Children asked if they could do the voices themselves (QC3) and emphasised that having a child’s voice throughout would be more relatable for other children (QC4). Parents/carers suggested that explaining the science around ADHD was important and should be presented in an easy and relatable way (QP6). Children also gave their ideas for how to visually present ADHD by providing descriptions of their own ADHD. For example, one child described it as colourful (QC5).

#### · Theme 3: Information about ADHD

Parents/carers highlighted the importance of basic information to help children understand what it means to have ADHD, including explaining the diagnosis and that it is not uncommon (QP7; QP8). Most agreed that the resource should include basic facts and figures about ADHD (QP9). Participants stated that there should be an emphasis on diversity and individual differences (QP10).

Children said that when they were diagnosed, they would have liked to know more about what ADHD is and what it means for them to have ADHD (QC6). They also agreed that it was important to include information such as the myths and facts (QC7).

#### · Theme 4: Self-management tips

There were discussions regarding the necessity to present self-management advice and tips for children to help with difficulties both at home and at school. This empowers the individual and family with tools that can help (QP11). The children shared their top tips when diagnosed with ADHD to others (QC8 & QC9).

Diversity and individual differences were highlighted, where not everything will work for everyone. Most agreed that it was important to try and be adaptive and flexible in different situations, and to let others know what is helpful or unhelpful for you (QP12).

#### · Theme 5: Positive aspects of ADHD

FG participants highlighted the need to talk about ADHD in a positive way, as this can boost self-esteem in newly diagnosed children. Families noted that the material currently available is mostly negative (QP13). Participants agreed that the advantages and strengths associated with having ADHD should be made clear and to show that ADHD has advantages (QP14). The children also shared positive features and what they think about their ADHD (QC10).

#### · Theme 6: Challenges of having ADHD

There was also agreement that having ADHD can be difficult (QP15). Children expressed some difficulties they face especially at school or with friends (QC11; QC12). Participants felt that there is a need to be aware of the challenges that can occur, so they can try to manage them (QP16). Some of the difficulties included co-occurring conditions such as autism and anxiety, difficulties with friendships, and social communication (QP17).

### Views of health professionals on themes – (stage 2)

In total, 23 health professionals completed questionnaires eliciting their opinions on the preliminary themes identified from the lived experience FGs. This included child and adolescent psychiatrists, specialist nurses/nurse practitioners, paediatricians, clinical psychologists and speech and language therapists, from a range of services (Generic Child and Adolescent Mental Health Service (CAMHS), Neurodevelopmental services, Complex Intervention and Treatment Teams (CITT), Community Paediatrics, academic psychiatrists). Overall, health professionals endorsed the preliminary themes that were generated from the first FG with children and parent/carers.

#### · Themes 1 & 2

Health professionals thought that a video would be useful to help raise awareness and reduce stigma. (quote 1, health professionals (QHP1), table 2). They also highlighted that there is no accessible and accurate information readily available for children with ADHD (QHP2). Professionals suggested presenting information in a way that appeals to children and to use visual aids to facilitate understanding (QHP3).

#### · Theme 3: Information about ADHD

Health professionals viewed this theme as particularly important to support children with ADHD to improve their understanding and acceptance of their condition (QHP4). They shared their experience that whilst helpful, sometimes analogies and abstract ideas can be confusing (QHP5).

#### · Theme 4: Self-management tips

Health professionals noted the significance of empowering children and parents, providing them with knowledge and tools that can help with self-management (QHP6) and how these strategies can be used alongside other interventions, such as medication (QHP7). They emphasised that the key is for children to get support to develop strategies as not everything works for everyone (QHP8).

#### · Theme 5: Positive aspects of ADHD

Most health professionals highlighted that misconceptions and stigma about ADHD mean that it is often portrayed negatively (QHP9). They emphasized focusing on positives and strengths especially as low self-esteem is a common issue among children with ADHD (QHP10).

#### · Theme 6 (Challenges of having ADHD)

In line with lived experience participants, health professionals agreed that it is important to set realistic expectations and understand that there are different challenges with having ADHD. Recognising and identifying one’s difficulties and challenges is essential and enables individuals to find ways to overcome the difficulties (QHP11; QHP12).

### Feedback on themes (stage 2)

During the second FG, participants agreed that the preliminary themes represented their views, rating them as important to very important. Similar points raised during the first FG were reiterated with some specific suggestions, for example, *‘be careful not to box ADHD up’* which reflects the sub-themes of diversity (theme 3) and comorbidity (theme 6) (see table 1). On the theme of positive aspects of ADHD (theme 5), it was important to convey the message that *‘ADHD is just one part of who you are’* and *‘it doesn’t stop you from being successful*.*‘* Participants wanted to avoid terms such as *‘superpowers’* which might create unnecessary high expectations but to stress that *‘it’s okay to struggle’* (theme 6). Parents also like the idea of the video including tips (theme 5) as *‘it’s hard for us parents to get the kids to take on board advice’*. There were also discussions on the format, colours, storyline and visual suggestions for the video (theme 2; Figure 3). The children and parents/carers agreed that the video should be told from a *‘child’s perspective with a child’s voice’* as they feel *‘children would be more receptive to hearing another child’* (theme 2).

**Figure 3:**
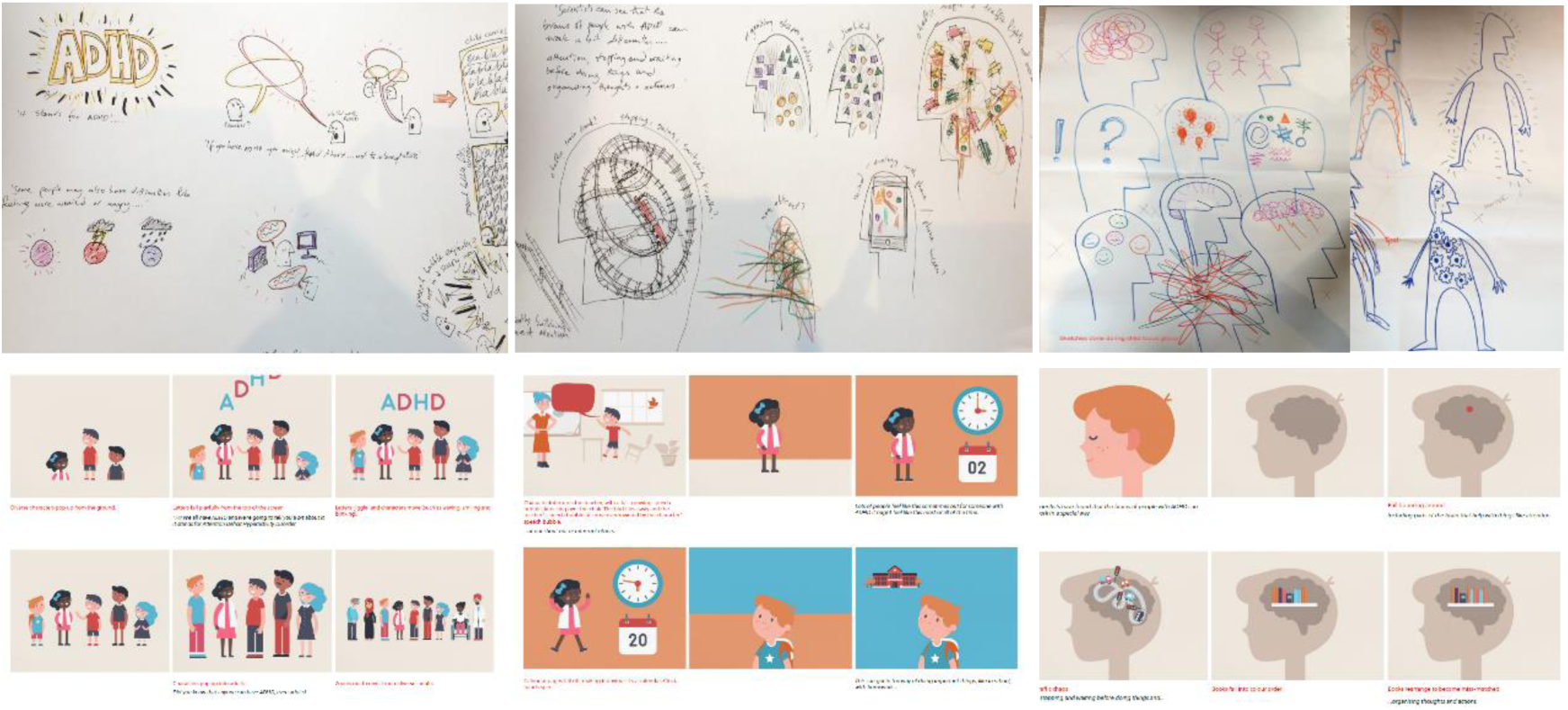
Development of visual ideas for the animation video: initial sketches (top 3 images) and screenshots from storyboard (bottom 3 images)

### Development and revision of the script, storyboard and animation (stage 3)

Based on these key themes and in line with scientific findings on ADHD, the script was carefully developed to ensure content was clear, developmentally appropriate, factual and without jargon. The script was phrased to a Flesch-Kincaid reading level of 7 (easy to read) (Farr et al., 1951). During the third FG, there were discussions of the language, diversity of characters and order of storyline. Children and parents/carers liked the characters and depictions of what is happening in the brain. Health professionals (n=5) agreed in individual discussions that the storyboard covered the core themes and that the information was clear and accurate. They liked that the animation was from a child’s perspective. They commented that the characters were warm, bright and cheery, which was appropriate for the target audience. Feedback and suggestions on improving the storyboard included addressing confusing images (e.g. illustration of DNA) and unhelpful stereotypes (e.g. doctor with a stethoscope) and increasing diversity of characters.

The research team reviewed the stakeholders feedback and shared them with the digital animation company. Once the final draft of the storyboard was agreed upon, the video was produced, with several edits to ensure that the video was clear, accurate, developmentally appropriate and included voiceovers from some participants (n=5).

### Animation video launch and feedback (stage 4)

The launch event was held in October to coincide with ‘ADHD awareness month’. Approximately 60 families attended the open day, and 70 families and professionals participated in the video animation screening. We received positive feedback (verbal and written feedback) from families/carers, for example noting pleasure: *“That we’re important enough for you to put this event on for us”* and around the *“Wealth of information from all three talks, great understanding with children and food”*

The response to the ADHD animation video was positive. Families/carers expressed that they liked *‘that children with ADHD took part’*, ‘*it was positive, informative and short enough for children’*, and *’I loved it, the diversity of characters and the balance of information in particular’*.

Health professionals described the video as having *‘Just the right amount of information’, ‘Visually appealing’, ‘sensitively worded’*, *‘A great animation – explained in a very clear child (and adult) friendly way’,* and *‘Loved the part about positive aspects’*.

To date, the ADHD animation video has received a positive response online and on social media. At the time of writing (30.05.2025), the ADHD animation video has reached around 639,000 views on YouTube.

## Discussion

The aim of this project was to co-develop and design a psychoeducation animation video about ADHD for children aged 7-11 years in partnership with children with ADHD and their parents/carers. This collaboration has led to the creation of a much-needed resource for the community, enhancing translation of research for societal benefit.

The online digital format reflects how children and young people (and many parents, carers and practitioners) obtain information (Bussing et al., 2012b; Ito-Jaeger et al., 2021). Design features suggested during the FGs such as including subtitles and child voiceovers, including diverse characters and colours was incorporated to help make the video more accessible and appealing to a wide range of viewers. Additionally, by adopting a bilingual approach, the animation video was more inclusive and has the potential to be translated into other languages. In line with the call for increasing participation of those with lived experience in research (Bisset et al., 2023), this project used a co-production approach with children with ADHD as well as their parents/carers and health professionals.

The content for the animation draws upon lived experience and perspective from a small group of children with ADHD and families from South Wales and therefore may not be representative of views of all those with ADHD. There are limited (qualitative) findings on content for psychoeducation from the perspective of those with lived experience for comparison, but the key themes derived from the FGs reflect some of those included in a systematic review of qualitative literature on the information and support needs of people with ADHD after diagnosis (NICE (UK), 2018). Most studies in this review focused on parent/carer views, and our project builds on these findings by including perspectives of children with ADHD and reiterating the importance of the themes identified.

Some of the themes highlighted in the review included the importance of being aware of the impact of ADHD and its association with other conditions/behaviours. This is reflected in the animation content themes 5 and 6, positive aspects and challenges of ADHD. In the NICE review, people wanted more information on ADHD upon diagnosis, and this was reflected around the theme of the need for the video and information about ADHD. Results from the review also highlighted that people with ADHD experience stigma, embarrassment and doubt about their diagnosis. This ties in with theme 5 where stakeholders expressed the importance of highlighting positive aspects of ADHD, stating that current resources they have found are mostly negative. A recent review recommends avoiding judgmental language that elicit negative stereotypes and over emphasis on challenges (Bisset 2023). This is important given that children and young people with ADHD and their families/carers experience low self-esteem and low confidence which results in self-stigma (Jelinkova et al., 2024; Sikirica et al., 2015).

The project has led to the co-production of a digital resource which is aimed to provide support and improve understanding of ADHD to children and their families/carers after being diagnosed with ADHD. Although it is aimed at children, it is hoped (and feedback from the launch event suggests) that it will help increase awareness and understanding about ADHD to the wider community. It also provides a proof of concept for a resource development framework for future co-production with a previously poorly represented group.

### Study limitations

It is possible that there was selection bias in that FG participants who volunteered may already have considered the need for a digital psychoeducation resource. Although we had a mixed range of children of different ages, more boys took part in the FGs. However, this may be representative as ADHD is more commonly diagnosed in males (Martin et al., 2024). Whilst participation from both parents was encouraged, parents/carers that took part were all female. Health professionals were recruited via researcher-established networks, although responses were obtained from different types of health professionals. The video animation does not provide a comprehensive guide to ADHD, but it includes the main themes highlighted in stakeholder discussions and is a good starting point for introducing ADHD. Additionally, the video may not be suitable for those with comorbidities such as learning difficulties, although the families of such children were involved in the process. Discussions with stakeholders generated more ideas about a much wider range of resources. However, there was only sufficient funding to develop just one resource within the given time frame.

### Conclusion

The completion and success of this project resulted in a free and widely accessible evidence-based digital resource for children newly diagnosed with ADHD and their families. The animation video is a valuable resource for families via the support groups and clinical services providing support and a better understanding of ADHD more widely. Future work in this area is to formally evaluate the ADHD animation and build on this framework to develop a more comprehensive support package for individuals diagnosed with ADHD and their families and carers.

The ADHD animation video is available on the National Centre for Mental Health (NCMH) website, https://www.ncmh.info/videos-and-podcasts/animations/, on the ACAMH website as a blog https://www.acamh.org/blog/lets-talk-about-adhd/ The ADHD animation video is available in both English (https://youtu.be/YeamHE6Kank) and Welsh (https://youtu.be/JbdD3kCZBVs). There is a postcard for dissemination at engagement events and clinics (Appendix 4)

## Data Availability

The data that support the findings of this study are available as excerpts in the manuscript (see table 2). The full data transcripts are not publicly available, even upon request, in line with ethical approval and consent obtained from participants.

## Acknowledgements

We would like to thank all the children, their families/carers, and health professionals in the field who shared their experiences and research to help us co-produce the animation. Our thanks to the Wellcome for funding (ISSF3 Public Engagement Proof-of-Concept Award) our project and for making the animation video possible, and to the digital media company (Spindogs) for their work.

## Key points and relevance

- There are not sufficient co-produced resources on ADHD readily available, especially for children with ADHD
- Aims are to connect research with public need, enhancing translation of research for societal benefit
- The outcome of project has led to an evidence-based bilingual resource for children newly diagnosed with ADHD
- Outlines the framework and proof of concept for future co-production with children with ADHD

