## Appendices for "Co-development of a digital animated video on ADHD with children and families/carers"

### Appendix 1 : Topic Guide – Children

| |
|---|
| **1a: Focus group 1 (children)**<br><br>Ground rules & Ice breaker session<br>• Do you think it is important to have this video<br>• How would you describe ADHD to me if I didn't know anything about it?<br>• Did you know what ADHD was before?<br>• Describe / draw what ADHD feels like to you<br>• What was it that helped you understand?<br>• How do you tell other people about ADHD? What does having ADHD mean: at school, with friends, for your family<br>• What do you think should be in the video?<br>• What do you think would help children understand what ADHD is?<br>• What should we call the video?<br>• Introduce facts and myths (displayed on posters around the room and on slides during discussion). Did you know about these before? Is it helpful to know this? Useful to be include in the video? Rate which one you think should be included |
| **1b: Focus group 2 (children)**<br><br>Discuss and further develop ideas from last session<br>• What ADHD is: how should we describe ADHD?<br>• Good things about my ADHD<br>• Having ADHD can be hard<br>• There are some things you might find difficult - Examples?<br>• Tips<br>• Discuss video design features<br>• Suggestions for a storyline for video |
| **1c: Focus group 3 (children)**<br><br>Setting: The storyboard was printed out as posters and stuck on poster boards. Children were instructed to go around the poster boards, putting a star sticker on bits of the storyboard that they liked and spotty stickers on bits that they didn't like. Children were accompanied by facilitators using the question below as a guide:<br>• What did you think of the characters / colours?<br>• Is there anything you didn't understand?<br>• Is there anything that you think is not right?<br>• Tell me one thing you do that helps with your ADHD?<br>• What did you like most about it?<br>• What did you not like about it? |

### Appendix 2 : Topic Guide – Parent/carers

#### 2a: Focus group 1 (parents/carers*)*

**Experiences of ADHD before diagnosis**

- Did you know anything about ADHD?
- What did you know about it?
- What did you think of ADHD?

**Experience upon receiving a diagnosis**

- What information did you receive when your child was first diagnosed?
- How was it explained to your child?
- What would you have liked to know?
- What do you think would be most useful for children to know?

**Experience of looking for information about ADHD**

- Have you tried looking for **videos**/information on ADHD?
- Was it helpful/useful?

**What do you think should be in this video?**

- What should we include?
- What should it be called?
- Should it be factual or in form of a story?

**What do you think of these myths and facts?**

- Did you know about this?
- Is it helpful to know this?
- Useful to include in video?

<u>Myths and facts about ADHD</u>

1. ADHD is not just 'naughty' behaviour
2. ADHD is not 'new'
3. ADHD hasn't become more common recently
4. Some children grow out of ADHD, but some don't
5. A diagnosis of ADHD doesn't necessarily mean a child will be prescribed medication
6. There's no test for ADHD
7. ADHD is certainly not caused by bad parenting or lack of discipline.
8. There is no single cause for ADHD
9. Genetics are important, but there's no 'ADHD gene'
10. ADHD can run in families, but doesn't always

#### *2b:* Focus group 2 (parents/carers*)*

Welcome back! We have now put together the information and ideas from both parent and child groups and picked the ones that were most popular. Please go through this questionnaire individually and we will then go through each question and discuss each one together.
(*Each theme was introduced separately, followed by the next three questions after each theme)

1. What do you think of this theme? Please explain why
   **Not important** **Somewhat important** **Important** **Very important**

2. Do you have any suggestions that should be included into this theme?

3. Is there anything related to this theme which you feel is unhelpful that we should avoid (e.g. analogies, tips, clichés)?

Based on all the themes discussed, are there any important areas or themes that have not been covered?

***2c:* Focus group 3 (parents/carers*)***

Setting: Storyboard printed out as poster and stuck on walls in seminar room. Participants went around with post it notes leaving feedback, comments and suggestions using the questions below as a guide.

1. To what extent do you feel that we have addressed the themes mentioned above?
2. Are there any messages that you feel are factually incorrect or not appropriate for the target group (children aged 7-11 with ADHD)? If yes, please let us know what this is
3. Are there any messages that you feel are poorly described? If yes, please let us know what this is
4. What did you like it about it?
5. Is there anything that you did not like? Do you have any suggestion for improvements?

### Appendix 3: Health professionals questionnaire

#### 3a: Health professionals questionnaire – Stage 2

This short questionnaire should take 10-15 minutes to complete and will ask for your input on each of the themes that have been suggested. Your responses will be anonymous and your contribution would be very much appreciated.

- Which service do you work in?
  Generic CAMHS / Paediatrics / Neurodevelopmental service / Other (please specify)
- What is your role? (e.g. Psychologist, Specialist Nurse or Therapist, Psychiatrist, Paediatrician)

(*Each theme was introduced separately, followed by the next three questions after each theme)

1. What do you think of this theme? Please explain why
   **Not important** **Somewhat important** **Important** **Very important**
2. Do you have any examples/analogies to explain ADHD to a child or family that you think are helpful or would like to share?
3. Is there anything related to this theme which you feel is unhelpful to children and families/carers that we should avoid (e.g. analogies, tips, clichés)

Based on all the themes discussed above, are there any important areas or themes that have not been covered?

#### 3b: Health professionals questionnaire – Stage 3

**Please let us know what you think of the ADHD animation storyboard.** This should take 5-10 minutes to complete.

1. To what extent do you feel that we have addressed the themes mentioned above?
   - All well addressed
   - All somewhat addressed
   - Some well addressed, some not
   - None well addressed
2. Are there any messages that you feel are factually incorrect? If yes, please let us know what this is
3. Are there any messages that you feel are poorly described? If yes, please let us know what this is

4. Are there any messages that you feel are not appropriate for the target group (children aged 7-11 years with ADHD)? If yes, please let us know what this is

5. What did you think of it overall?

6. What did you like it about it?

7. Is there anything that you did not like? Do you have any suggestion on how we can improve it?
For example: we realise that the picture of the doctor taking the boys heart rate whilst placed on a high trolley is not really a good representation. We will suggest that this is changed.

Appendix 4: Postcard on ADHD animation for dissemination in clinics and events (postcard front- left, postcard back-right)

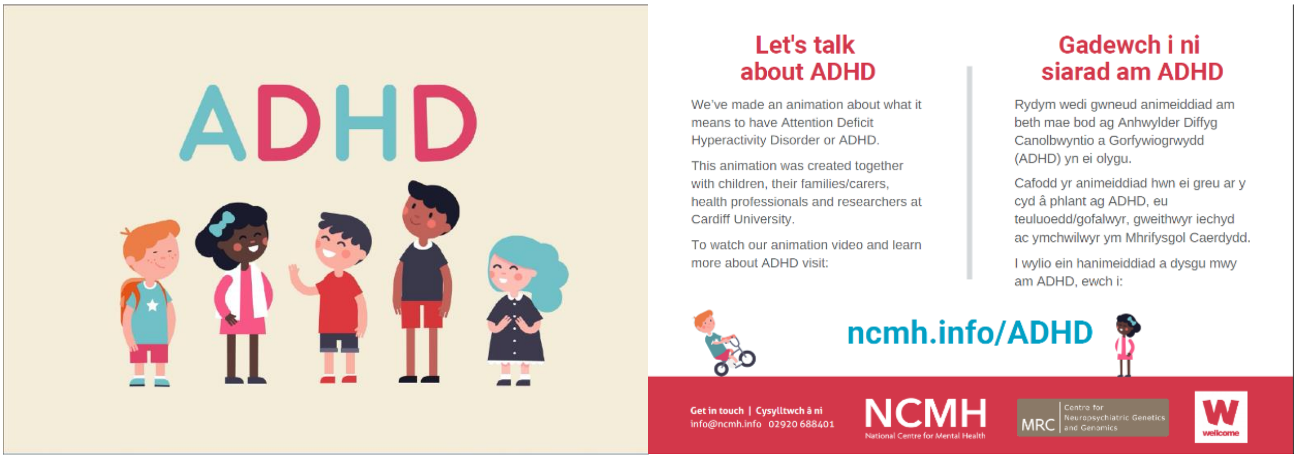
